# Strength Training Exercises to minimise late Effects of childhood Leukaemia or lymphoma among adolescents – The STEEL study – protocol for a national multicentre randomised controlled trial

**DOI:** 10.1101/2025.08.28.25334630

**Authors:** Henrik Riel, Mathias Vedsø Kristiansen, Birgitte Klug Albertsen, Martin Kaj Fridh, Christina Friis Jensen, Hanne Bækgaard Larsen, Mathias Rathe, Stine Svilshave, Ruta Tuckuviene, Clara Vad, Joachim Wiskemann, Pascal Madeleine

## Abstract

**Background:** Adolescent cancer survivors (ACS) often experience long-term impairments in muscle strength, physical function, and quality of life. Strength training may help address these deficits, but there is limited evidence from randomised trials. This study investigates the efficacy of progressive strength training compared to low-load circuit training in increasing muscle strength over 16 weeks among ACS.

**Methods:** In this multicentre randomised controlled trial, we will recruit 60 ACS successfully treated for acute lymphoblastic leukaemia or lymphoma. Participants aged 10–19 years are recruited from paediatric oncology departments across Denmark. They are randomised stratified by sex to either: 1. a 16-week partly supervised progressive strength training programme (STEEL) with individually tailored exercises targeting major muscle groups, progressing in load and volume over time; or 2. a 16-week partly supervised circuit training programme involving aerobic and low-load strength exercises. The primary outcomes are the change in isometric muscle strength in elbow flexion and extension, and knee flexion and extension from baseline to the 16-week follow-up. Secondary outcomes include physical function (dynamic strength, rate of force development, grip strength, and walking endurance), health-related quality of life, metabolic markers, bone mineral density, movement-evoked pain, exercise compliance and fidelity, physical activity level, and participant satisfaction.

**Discussion:** This trial addresses the need for evidence-based rehabilitation strategies in ACS. It is designed to detect short-term effects of supervised strength training on key physical health outcomes using a robust, blinded, and clinically relevant comparison. Results may inform future survivorship care and guidelines.

**Trial Registration:** ClinicalTrials.gov identifier: NCT05673152. Registered January 4, 2023.

## Background

In Denmark, roughly 200 children and adolescents are diagnosed with cancer each year. Fortunately, treatment has improved over the years, and now more than four in five children are long-term survivors compared to less than half the number 30 years ago.(1,2) With an increasing number of survivors, there is an increasing demand for the rehabilitation of children, as surviving is not synonymous with being fully recovered. Childhood cancer survivors have a high risk of treatment-related health complications that continue into adulthood, including type 2 diabetes, insulin resistance, dyslipidaemia, obesity, heart disease, low muscle strength, reduced physical function, and reduced bone mineral density.(3–5)

One of the side effects of cancer treatment, including chemotherapy, is a decline in muscular strength. This decline often emerges early in the course of the disease and may persist beyond the completion of therapy. Reduced muscular strength can impair adolescents’ ability to engage in physical activity, potentially leading to long-term adverse outcomes (4,6,7). Most studies investigating the effects of physical training for patients during cancer treatment have focused on performing nonspecific exercises involving, for instance, motor coordination (8) However, the evidence supporting exercise interventions in adolescents with cancer is less robust than in adults (8) This may, in part, reflect limited evidence is mostly due to methodological limitations in existing with poorly designed studies, (e.g., small sample sizes and short duration), but it may also be due to low exercise compliance (9) It has been suggested that introducing exercises when chemotherapy is ongoing may hamper compliance, as both children and their parents may be too affected physically and mentally to be able to perform exercises. Furthermore, recruiting participants during cancer treatment for exercise interventions may be challenging, as parents may not be able to assess the requirements of being involved in the study or have the mental capacity necessary.(9) This is further corroborated by the general lack of knowledge concerning the safety and efficacy of strength training in this population, as well as the limited knowledge concerning exercise prescriptions.(10) In a recent systematic review, Vasilopoulou et al.(11) have reported that interventions promoting physical activity are feasible and well-accepted among adolescent cancer survivors (ASC). A recent prospective randomised trial has shown that adaptive variable strength training ameliorates strength and muscle thickness.(12) A recent study among children treated with chemo-/radiotherapy in a Danish setting had a high acceptance rate of exercise and found promising results on cardiorespiratory fitness following a physical activity program.(13) Despite the superiority being sustained one year after treatment compared with the control group, the ACS still had a lower cardiorespiratory fitness compared to the community control.(14) Hence, there is a need for studies that investigate the effects of strength training among ACS who suffer from late effects after treatment because they did not have the desired effects of exercise or were unable to perform exercises when cancer treatment was ongoing.

Many previous studies investigating the effects of exercise to increase muscle strength and muscle mass have not tailored exercises to do so or have not reported the exercise programme sufficiently to allow for replication.(8,9,15) Adolescents may increase strength from resistance training beyond maturation and growth, provided that the exercises are of sufficient volume, intensity and duration.(16) Yet, studies thus far have primarily used low-load activities such as circuit training and aerobic exercises, which do not contribute to an increase in muscle strength and muscle mass.(8) A strength training protocol with a sufficiently high intensity is considered much more ideal when the goal is to increase muscle strength, muscle mass, and bone mineral density, which should be the goal considering the deficiencies ASC have.(16) Before adequately investigating heavy strength training among this population, it remains unknown if the same benefits of sufficient strength training among healthy adolescents are evident among adolescents who have survived cancer and if this may lead to a more physically active lifestyle. To date, there is very little knowledge about the dosage of strength training for cancer survivors.(17) Therefore, this study aims to investigate the efficacy of STEEL strength training compared to low-load circuit training in increasing strength over 16 weeks among adolescents who have survived childhood leukaemia or lymphoma. We hypothesise that STEEL will be superior to circuit training in increasing strength among ASC after 16 weeks.

## Methods

### Study design

The study is designed as a national multicentre randomised controlled study. Participants are randomised (1:1) to either: 1) STEEL strength training, or 2) circuit training. The study is conducted at Aalborg University, Copenhagen University Hospital, Odense University Hospital, and Aarhus University Hospital and was designed in collaboration with parents of childhood cancer survivors and an adult childhood cancer survivor suffering from late effects. Participants must attend three examinations at their respective hospitals or the university: baseline, 8 and 16 weeks after the intervention starts. Future reporting of the trial will follow CONSORT guidelines for reporting non-pharmacologic treatments.(18) Reporting of this protocol follows the SPIRIT statement.(19) The trial was planned in accordance with the PREPARE Trial guide.(20) The trial is being conducted according to the Declaration of Helsinki III, and the protocol, template informed consent forms, and participant information were approved by the Ethics Committee of North Denmark Region (N-20230055). All participants provide informed consent before enrolment. When participants or their parents/legal guardians sign the consent form, they agree that they have been adequately informed about the purpose, methods, advantages, and disadvantages of participation; the participants know that participation is voluntary; and they can withdraw from the trial without losing their current or future rights to receive treatment. Before the inclusion of the first participant, the trial was registered on <u>clinicaltrials.gov</u> (NCT06299722).

### Roles and responsibilities

The project manager is a physiotherapist with 14 years of experience treating patients with musculoskeletal conditions and 10 years of experience conducting research with exercise interventions. He is responsible for managing the project, collecting data among some of the participants at Aalborg University, training data collectors, and the physiotherapists and trainers supervising the exercises, and conducting the statistical analyses. Each study site has a medical doctor responsible for determining participant eligibility based on medical records and a physical examination. Data collectors include physiotherapists, medical students, nurses, sports scientists and clinical researchers. Before data collection, all received a minimum of 1 hour of training from the project manager. Half of the training sessions are supervised by either physiotherapists or certified trainers who receive a 30-minute online induction training by the project manager in which they are informed about the trial’s procedures, background, and the two exercise protocols.

### Recruitment

A participating researcher from each hospital contacts potential participants who have been treated for cancer at Aalborg University Hospital, Aarhus University Hospital, Odense University Hospital or Copenhagen University Hospital by searching medical records and internal registries. After identifying potentially eligible participants, an invitation is sent via e-mail to those identified at Aalborg University Hospital or Aarhus University Hospital, including information about the trial. The potential participants or parents/legal guardians (for those younger than 18 years of age) are then contacted by telephone afterwards. At Copenhagen University Hospital and Odense University Hospital, potentially eligible participants are contacted by phone or during scheduled checkups.

### Eligibility criteria

We recruit adolescents aged 10 to 19 years who have been treated for acute lymphoblastic leukaemia or lymphoma (Hodgkin’s or non-Hodgkin’s). Inclusion criteria include: (1) 10 to 19 years of age at the point of inclusion; (2) a minimum of 12 months since the last chemotherapy with no upper limit; and (3) ability to understand the physical intervention and general participant advice. Exclusion criteria are: (1) participation in another research study that includes similar treatment; (2) pregnancy; (3) cardiac arrhythmia during exercise; (4) psychological disorders interfering with treatment, (5) presence of a clinical condition that needs immediate treatment; (6) planned surgeries within the subsequent 12 months that may interfere with performing exercises; or (7) any contraindications to performing physical exercise as evaluated by the recruiting medical doctor.

### Interventions

In both groups, exercises are performed twice a week, with a minimum of 48 hours apart from each other over 16 weeks. A physiotherapist or other certified trainer supervises one weekly session, whereas the other is performed unsupervised. An overview of the two protocols can be seen in Table 1.

**Table 1:** Overview of the exercise descriptors of the exercise protocols.

|  |  |  |  |
| --- | --- | --- | --- |
| **STEEL** |  |  |  |
|  | **Weeks 1 to 5** | **Weeks 6 to 10** | **Weeks 11 to 16** |
| **Load (RM)** | 12 | 10 | 8 |
| **Repetitions** | 10 | 8 | 6 |
| **Sets** | 3 | 4 | 4 |
| **Rest between sets (min)** | 1.5 | 2 | 2.5 |
| **Time under tension (eccentric-isometric-concentric-isometric) (s)** | 2-1-X-1 | 2-1-X-1 | 2-1-X-1 |
| **Circuit training** |  |  |  |

|  | Weeks 1-5 | Weeks 6-10 | Weeks 11-16 |
| --- | --- | --- | --- |
| **Rate of perceived exertion Borg scale (6-20)** | 14-15 | 16-17 | 18-20 |
| **Rounds** | 3 | 3 | 3 |
| **Time (s)** | 45 | 45 | 45 |
| **Rest between exercises (s)** | 15 | 15 | 15 |
| **Rest between rounds (min)** | 2 | 2 | 2 |
| RM: Repetition maximum |  |  |  |

#### STEEL strength training

The STEEL strength training consists of progressive periodised strength training, in which the intensity is set to fit the individual’s starting level and then gradually increases. During Week 1 to Week 5, the relative load corresponds to a 12-repetition maximum (RM). During Week 5 to Week 10, the relative load is 10 RM, and during Week 11 to Week 16, the relative load is 8 RM. The exercises are five compound movements that activate the large muscle groups of the legs, back, arms and torso. The physiotherapist selects one exercise for the lower extremities (e.g., squat or leg press) and four exercises for the upper extremities; one horizontal push (e.g., chest press), one vertical push (e.g., shoulder press), one horizontal pull (e.g., seated row), and one vertical pull (e.g., lateral pulldown) from an exercise catalogue to match the participant’s starting level. The exercises can be changed over time as long as the programme still includes five exercises in the aforementioned movement directions.

#### Circuit training

The circuit training is performed similarly to the circuit training employed in Braam et al. (21), using weight-bearing exercises with balls, hoops, and running activities included. There are 10 stations that include strengthening and cardiovascular exercises, such as squats, hopping, abdominal crunches, stationary running, front-lying swimming, ski jumps, push-ups, ball throws, ring pulls, and hopscotch. Each station lasts 45 seconds, and there is a 15-second break between each station. The circuit is repeated for three rounds. To ensure progression in intensity similar to that of Braam et al. 2018, we progress the participants’ rating of perceived exertion (RPE) over time. During Week 1 to Week 5, the RPE during exercises will be 6-7. During Week 5 to Week 10, the RPE will be 8-9, and during Week 11 to Week 16, the RPE will be 10. To accommodate the preference of the patient group, which has previously been investigated, the training sessions are carried out at well-equipped local training facilities such as fitness centres or physiotherapy clinics within a 30 km radius of participants’ homes and, if possible, together with either a friend the adolescent may bring along or other trial participants.(22)

During baseline, we provide the participants from both groups and parents with an educational session with information regarding lifestyle, including nutrition, sleep, and risks of late effects. We also provide emotional support and information about expected exercise-related muscle soreness and encourage them to be physically active. They are allowed to participate in sports as much as possible, but are asked to refrain from performing strengthening exercises other than those included in the trial.

### Randomisation and blinding

Participants are stratified by sex and block randomised (block sizes of 2 to 6) at 1:1 to either STEEL or the circuit training programme. A researcher not involved in the trial generated the allocation sequence using a random number generator (www.sealedenvelope.com) and is the only person to know the block sizes. The project manager trained the researcher in generating allocation sequences, and the process was piloted before they generated the trial’s allocation sequence. The researcher placed notes with the randomisation into consecutively numbered opaque envelopes kept in a locked cabinet. After a participant has been included and all baseline measurements have been taken, the project manager opens the next envelope and assigns the participant to STEEL or the circuit training.

Besides at Aalborg University, the data collectors at each study site are blinded to the allocation. As the project manager collects data at Aalborg University, blinding is not possible.

### Variables

The outcome measures and measurements are summarised in Table 2.

**Table 2:**
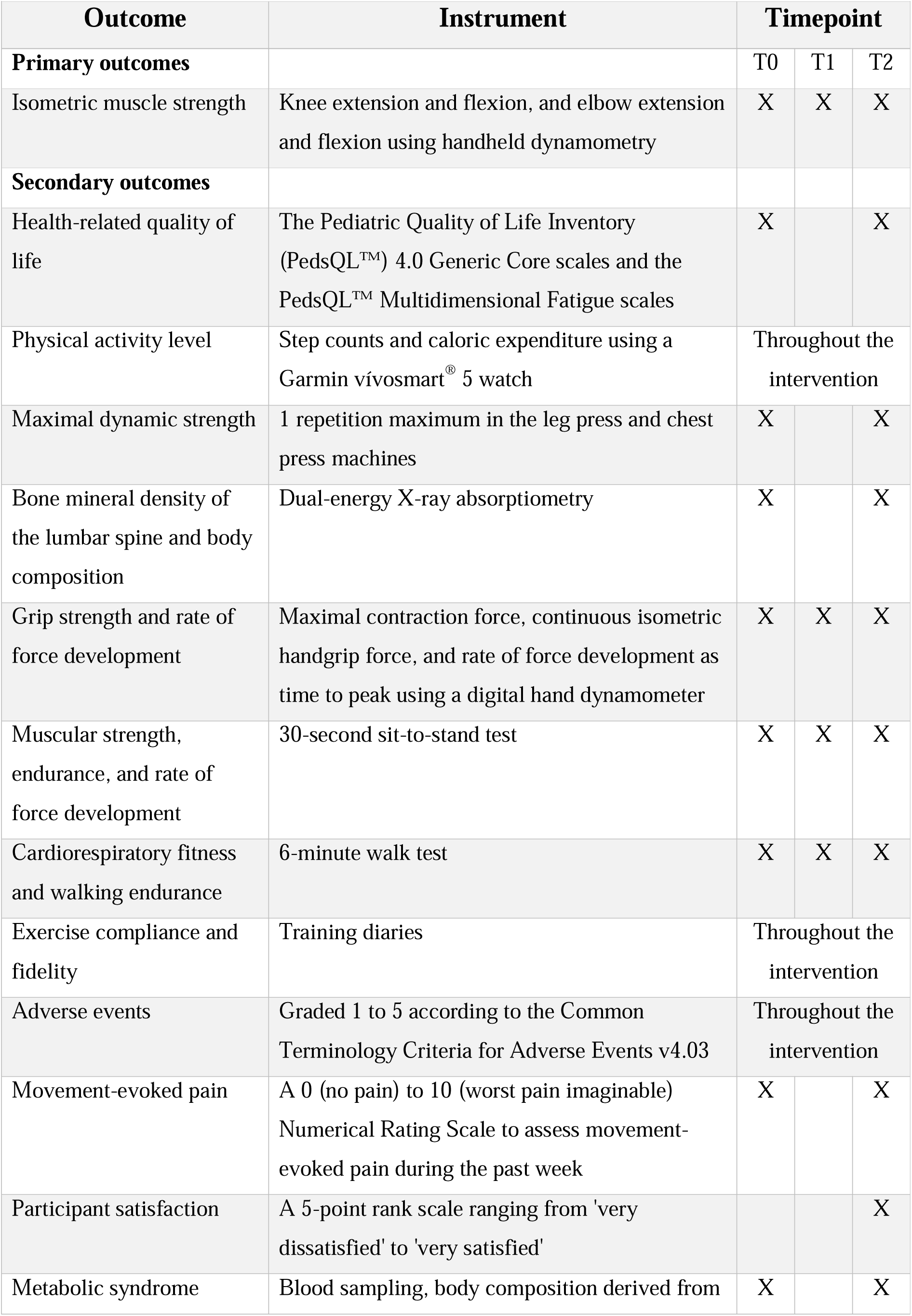
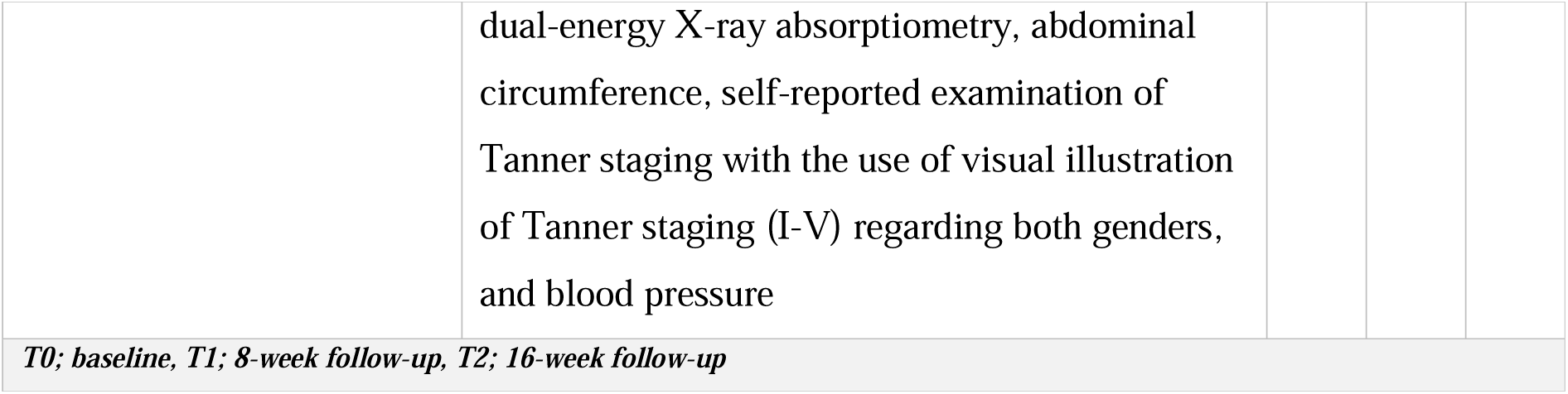
Assessments and instruments.

#### Descriptive

During baseline, we collect the following participant characteristics: age, sex, ethnicity, menarche, cancer diagnosis (leukaemia or lymphoma), treatment protocol, relapse (yes/no), age at diagnosis, time since last chemotherapy, late effects, alcohol consumption, smoking status, medication use, known illnesses, and participation in sports or recreational physical activities.

#### Primary outcomes

The primary outcomes are changes in isometric strength measured in Newton (N) based on tests of knee extension and flexion, and elbow extension and flexion using handheld dynamometry from baseline to the 16-week follow-up (23). All tests are performed twice in each movement direction, and the average of the two tests will be used. Before the tests, participants perform two trial tests in which they only exert 50% of their maximum force to familiarise themselves with the test. Next, they perform two maximal contractions for 10 seconds separated by a 1-minute rest. During the elbow extension and flexion tests, the participant lies supine with the elbow at a 90-degree angle. During the knee extension and flexion tests, the participant is seated with the knee and hip at a 90-degree angle. The tests are performed during baseline and the 8-week, and the 16-week follow-ups.

#### Secondary outcomes

The secondary outcomes include:

##### 1. Health-related quality of life

We use the Danish version of the Pediatric Quality of Life Inventory (PedsQL™) 4.0 Generic Core scales and the PedsQL™ Multidimensional Fatigue scales (8,15) to assess health-related quality of life. The PedsQL 4.0 Generic Core is a brief 23-item measurement model that evaluates quality of life in four areas: physical, emotional, social and school functioning. The questionnaire scores range from 0 to 100, and higher scores indicate a better quality of life. The PedsQL™ Multidimensional

Fatigue includes 18 fatigue-related questions and uses the same scoring as the PedsQL 4.0 Generic Core. The questionnaires are filled out during baseline and the 16-week follow-up.

##### 2. Physical activity level

We will assess participants’ step counts and caloric expenditure throughout the intervention using a Garmin vívosmart^®^ 5 watch (Garmin Ltd., Kansas, USA). Participants are given the watch during baseline and are asked to wear the watch as often as possible. Only days with recorded activity will be included in future analyses. At the 16-week follow-up, daily step counts and caloric expenditure during the 16 weeks are extracted.

##### 3. Maximal dynamic strength

During the first supervised training session, the physiotherapist or trainer tests the one repetition maximum (1RM) in the leg press and chest press machines at the training facility. The test will be repeated during the final supervised training session, and any change is reported as the relative increase in the 1RM. The test begins with a general warm-up using a light load (∼40–60% of estimated 1RM) for several repetitions. Following a 1-minute rest, the load is increased to an estimated 3–5 repetition maximum (RM), and the participant completes 1–3 repetitions. After a 2-minute rest, the load is increased to an estimated 2–3RM, followed by 1–2 repetitions. A rest period of 2–4 minutes is then provided. Subsequently, the load is increased by 5–10% for the chest press and 10–20% for the leg press, and the participant attempts a single repetition (1RM attempt). If successful, the load is further increased by the same increment after a 2–4 minute rest. If the attempt is unsuccessful, the load is reduced by 2.5–5% for the chest press or 5–10% for the leg press, and a new attempt is made following a 2–4 minute rest. This process continues until the highest load that can be lifted once with proper form is identified.

For the chest press, machine and seat height are adjusted so that the handles align with the mid-sternum. Participants are instructed to keep their back and head in contact with the backrest throughout the movement. A valid repetition requires a controlled movement through the full range of motion with near-complete elbow extension.

For the leg press, participants are positioned with knees flexed to approximately 90 degrees and the back and head resting against the back support. If available, participants are instructed to hold the hand grips. A valid repetition requires full range of motion and near-complete knee extension while maintaining proper posture.

##### 4. Bone mineral density of the lumbar spine and body composition

We measure the bone mineral density of the lumbar spine (L1-L4), calculated as the Z-score and body composition (%body fat, and lean body mass) by a dual-energy X-ray absorptiometry scanner located at each hospital or university.(4,9,15) The scan is performed during baseline and at the 16-week follow-up.

##### 5. Grip strength and rate of force development

Using a digital hand dynamometer, we measure the maximal contraction force in Newton (N) and continuous isometric handgrip force in seconds and rate of force development as time (in seconds) to peak. The participants use their dominant hand and are instructed to grip the hand dynamometer as quickly as possible and with as much force as possible. During the tests, participants are seated in a chair holding the dynamometer with their palms facing medially, and elbows flexed at 90°. The test is repeated three times, with each trial lasting 3 seconds. Second, a single fatiguing isometric trial is conducted where the participant maintains a grip corresponding to approx. 30% of the maximal voluntary contraction for as long as possible.(24) The test is performed during baseline and after 8 and 16 weeks.

##### 6. Muscular strength, endurance, and rate of force development

We use a 30-second sit-to-stand test to evaluate the lower extremities’ strength, endurance, and rate of force development. The participant sits on a chair with no armrests and is asked to stand up, reach full extension, and sit down as many times as possible within 30 seconds with their hands across their chest. The score is the number of repetitions performed during the test.(25) The test is performed during baseline and after 8 and 16 weeks.

##### 7. Cardiorespiratory fitness and endurance

We perform a 6-minute walk test to evaluate cardiorespiratory fitness and walking endurance.(26) Two markers are placed 20 meters apart, and participants are instructed to walk as far as they can within 6 minutes whilst the instructor provides standardised encouragement. They are not allowed to run or jog. The test is performed during baseline and after 8 and 16 weeks.

##### 8. Exercise compliance and fidelity

Exercise compliance and fidelity will be measured using training diaries, which the participants fill out themselves after the unsupervised training sessions and by the physiotherapist during the supervised training sessions. Exercise compliance relates to whether the training sessions have been performed, and fidelity relates to whether the exercises have been performed as prescribed regarding the number of repetitions, sets, and intensity. It is expressed as a percentage of the prescribed exercise.

##### 9. Adverse events

Adverse events will be collected throughout the trial and graded 1 to 5 according to the Common Terminology Criteria for Adverse Events v4.03. Participants are asked to contact the responsible clinician at the hospital where they were enrolled as soon as they experience any adverse event.

##### 10. Movement-evoked pain

We use a 0 (no pain) to 10 (worst pain imaginable) Numerical Rating Scale to assess movement-evoked pain during the past week. Participants answer the question during baseline and after 16 weeks.

##### 11. Participant satisfaction

We assess participant satisfaction with their respective intervention using a 5-point rank scale ranging from ‘very dissatisfied’ to ‘very satisfied’. Responses are dichotomised into “Satisfied” (a rating of “Satisfied” or “Very satisfied”) or “Dissatisfied” (a rating from “Very dissatisfied” to “Either/or”). This outcome is collected during the 16-week follow-up.

##### 12. Blood sampling

Blood samples are used to analyse the metabolic status of each participant following childhood cancer. Furthermore, we will use blood samples to evaluate the effect of strength and circuit training on cardiometabolic health. Based on the biochemical analysis of the blood samples, we analyse the following outcomes that relate to metabolic syndrome: (1) glucose, (2) glycated haemoglobin (Hba1c), (3) insulin, (4) proinsulin c-peptide, (5) total cholesterol, (6) high-density lipoprotein cholesterol, (7) low-density lipoprotein cholesterol, (8) very low-density lipoprotein cholesterol, (9) triglycerides, (10) glucagon, and (11) leptin. Furthermore, the Homeostatic Model assessment for Insulin resistance score is calculated as fasting plasma glucose (mmol/L) x fasting plasma glucose ((μU/L)/22.5) to estimate β-cell function (HOMA-B) and insulin resistance (HOMA-IR2).(27) The participants are asked to fast for 6 hours before blood sampling. For each participant, we will use 50 ml or max 1 ml/kg. Blood samples are collected during baseline and the 16-week follow-up.

##### 13. Metabolic markers

To assess markers of metabolic syndrome, we use the following measurements: (1) anthropometrics with body mass index (BMI) for adolescents aged 18 or 19 years and BMI standard deviation (SD) scores for children aged 10 to 17 years based on national reference material, (2) body composition including lean body mass and fat mass, and android/gynoid fat distribution assessed by dual-energy X-ray absorptiometry (DXA) adjusted for sex and pubertal stage, (3) abdominal circumference, (4) self-reported examination of Tanner staging with the use of visual illustration of Tanner staging (I-V) regarding both genders and (5) blood pressure. Measurements are made during baseline and at the 16-week follow-up.

### Sample size

Based on a previous study by Braam et al. using a physical exercise intervention among ASC, it may be expected to see a relative change in strength of at least 10%, though we hypothesise to see an even larger improvement after STEEL.(15) We will need to recruit 54 participants to detect a medium effect size of 0.6 between the two groups at 16 weeks with an alpha of 0.05 and a power of 80%. To account for a potential 10% dropout, we aim to recruit 60 participants.

### Statistical analyses

The primary analysis will be a linear mixed effects model to test between-group differences in strength measured by the handheld dynamometers with the participant as random effect. The baseline value, time (8 and 16 weeks), group allocation (STEEL or circuit training) and term for interaction between time and group are treated as fixed effect variables. We will employ the same analysis to explore other differences in the continuous outcomes. Effect sizes (e.g., standardised mean differences) will also be calculated to quantify the magnitude of between-group differences in continuous outcomes. Potential between-group differences in the number of training sessions performed, exercise fidelity, step counts and caloric expenditure are investigated using unpaired t-tests. To explore between-group differences in participant satisfaction and the number of adverse events, we calculate the risk difference, and the number needed to treat as 1/risk difference.

### Data monitoring and quality assurance

All data is stored electronically and managed in compliance with the General Data Protection Regulation (GDPR). The Danish Data Protection Agency may conduct unannounced inspections to ensure data safety. Participant data are entered into REDCap, while data processor agreements, collaboration agreements between Aalborg University and the regional hospital authorities, and study protocols are stored securely on Aalborg University’s internal server. To minimise data entry errors, data collection instruments in REDCap are designed to require completion of mandatory fields and to include field validation (e.g., an error message appears if a date field is completed in an incorrect format). The project manager reviews the data to ensure completeness and accuracy. In accordance with the European Code of Conduct for Research Integrity, all data will be retained for 10 years following completion of the trial.

## Discussion

This trial evaluates the effects of 16 weeks of semi-supervised, progressive strength training (STEEL) compared to circuit training on muscle strength, physical function, health-related quality of life, metabolic markers, and satisfaction in adolescent survivors of childhood acute lymphoblastic leukaemia or lymphoma. The trial addresses a critical need for evidence-based exercise interventions tailored to this population, who often experience long-term impairments in strength and physical function due to cancer and its treatment, as highlighted in a recent systematic review.(11)

Several design strengths enhance the robustness of the trial. Both interventions are delivered in person by trained physiotherapists or trainers, ensuring support and continuous progression, with half of the training session being supervised. Using a randomised, multicentre design with stratification by sex and assessor blinding reduces the risk of bias and increases the generalizability of findings across different settings. Outcome measures were selected based on their relevance to childhood cancer survivors and their sensitivity to change, particularly muscle strength, a key concern in this population.

The study also has some limitations. First, it does not include follow-up beyond the 16-week intervention period. Although the trial is designed to detect between-group differences in the short term, longer-term follow-up would be needed to assess the sustainability of intervention effects.

Second, blinding is only partial; participants and training personnel know group allocation. However, outcome assessors at three of the four study sites remain blinded to minimise detection bias. Finally, while participants are recruited from national paediatric oncology centres, those with higher symptom burden or logistical challenges may be less likely to participate, potentially limiting generalizability to the broader childhood cancer survivor population.

If the intervention proves effective, the findings may support the integration of progressive strength training into follow-up care for adolescent survivors of childhood cancer. This could help address common long-term treatment effects, such as muscle weakness and reduced physical functioning and inform future clinical guidelines for survivorship care.(3–5)

## Data Availability

All data produced in the present study will be available upon reasonable request to the authors

## Notes

### Competing Interest Statement

The authors have declared no competing interest.

### Clinical Trial

NCT06299722

### Funding Statement

This study was funded by The Independent Research Fund Denmark and the Danish Childhood Cancer Foundation

### Author Declarations

The Ethics Committee of North Denmark Region gave ethical approval for this work (N-20230055).

